# C-reactive protein and lactate dehydrogenase as prognostic indicators in COVID-2019 outpatients

**DOI:** 10.1101/2021.11.17.21265553

**Authors:** Keiko Suzuki, Takaya Ichikawa, Satoshi Suzuki, Yoko Tanino, Yasutaka Kakinoki

## Abstract

**Background:** It is critical for clinicians seeing outpatients with coronavirus disease 2019 (COVID-19) to identify those who will require oxygen therapy after the hospital visit. Although studies on biomarkers predicting mortality or ventilator requirement in hospitalized patients with COVID-19 have been conducted, research on biomarkers predicting the need for oxygen therapy in outpatients is sparse.

**Methods:** Patients with COVID-19 who visited Asahikawa City Hospital on an outpatient basis were included in the study. In total, 287 new outpatients visited between April 2021 and September 2021, and 142 underwent blood testing. All blood tests were performed before any treatments for COVID-19 were started. Demographic information, laboratory data, and clinical treatment information were extracted from the electronic medical records. Risk factors associated with oxygen therapy were explored.

**Results:** In total, 40 of 142 patients who underwent blood testing required oxygen therapy within 7 days after blood samples were taken, and all other patients recovered without oxygen therapy. C-reactive protein (CRP) and lactate dehydrogenase (LDH) levels were significantly higher in patients who required oxygen therapy, and their cutoffs were 36 mg/L (sensitivity, 0.802; specificity, 0.725) and 267 U/L (sensitivity, 0.713; specificity, 0.750), respectively. Multivariate logistic regression identified age, body mass index, CRP ≥ 36 mg/L, and LDH ≥ 267 U/L as significant risk factors for oxygen therapy requirement. This study suggests that elevated CRP and LDH levels are independent risk factors for oxygen therapy in outpatients with COVID-19. Further confirmatory studies are needed.

## Introduction

Biomarkers can play a key role in decision making by physicians in daily practice for patients with coronavirus disease 2019 (COVID-19) (Tabassum et al., 2021). It is important for clinicians to identify patients requiring hospitalization. To ensure the efficient allocation of medical resources, patients who require oxygen therapy should be prioritized for hospitalization (Yamada et al., 2021).

Vahey et al. reported that chronic hypoxemic respiratory failure with a requirement for oxygen supplementation, opioid use, metabolic syndrome, obesity, age ≥ 65 years, hypertension, arrhythmia, and male sex were significantly associated with the risk of hospitalization among patients with COVID-19 (Vahey et al., 2021). Yamada et al. created a clinical score for evaluating the need for hospitalization based on age, sex (male), BMI, medical history, and symptoms (Yamada et al., 2021). Although such information is helpful, it is sometimes difficult to precisely predict the prognosis of COVID-19 in daily practice without biomarkers.

Biomarkers of inflammation and coagulopathy can aid in identifying patients hospitalized with COVID-19 who are at risk for clinical deterioration (Anyan et al., 2020). Poudel et al. reported that the d-dimer level on admission is an accurate biomarker predicting mortality in patients with COVID-19 (1.5 µg/mL is the optimal cutoff) (Poudel et al., 2021). Li et al. reported that an elevated lactate dehydrogenase (LDH) level at admission is an independent risk factor for the severity and mortality of COVID-19 (Li et al., 2020). Rahman et al. reported that C-reactive protein (CRP), d-dimer, and ferritin levels at hospital admission represent simple assessment factors for COVID-19 severity (Rahman et al., 2021). The neutrophil-to-lymphocyte ratio (NLR) is also mentioned to be a strong predictor of the prognosis for patients with severe COVID-19 using a cutoff of 4.4 (Kilercik et al., 2021). Procalcitonin may also be useful for predicting prognosis (Hu et al., 2021). Decreased albumin and globulin levels were independent factors related to oxygen therapy in patients younger than 65 years who were admitted for COVID-19 (Ni et al., 2021). However, these studies used the data of patients who had been admitted to the hospital. Because the biomarkers’ level change during the stages of COVID-19, there appears to be no consensus regarding the biomarkers predictive of prognosis among outpatients who require admission and oxygen therapy.

Therefore, this study analyzed the data of outpatients to identify the biomarkers predictive of the need for oxygen therapy in patients with COVID-19. Additionally, we discussed the optimal cutoffs of the predictive biomarkers.

## Materials & Methods

### Study design and participants

From April 2021 to September 2021, outpatients newly diagnosed with COVID-19 who visited Asahikawa City Hospital as new patients were enrolled in the study. All patients were diagnosed by polymerase chain reaction for severe acute respiratory syndrome coronavirus 2 (SARS-CoV-2) genes and/or antigen quantitation testing.

The endpoint of this study was the requirement for oxygen therapy, which was definitely administered to patients with percutaneous oxygen saturation levels of ≤92%.

### Ethics

This study was approved by the ethics committee of Asahikawa City Hospital and conducted in accordance with the ethical standards set forth in the Declaration of Helsinki (1983). The opt-out method was used to obtain patient consent in this study.

### Data collection

All demographic, clinical, and outcome data were retrospectively extracted from the Asahikawa City Hospital record files. The demographic characteristics of the patients (age, sex, body mass index [BMI]); white blood cell count (WBC); NLR; LDH, aspartate aminotransferase (AST), alanine aminotransferase (ALT), d-dimer, and ferritin levels; comorbidities, and clinical courses were recorded. The time from the onset of symptoms to blood sampling was also recorded. All blood samples were taken before patients received any treatment for COVID-19.

### Statistical analysis

Continuous data were analyzed using the Mann–Whitney U test and presented as the mean (interquartile range). For categorical variables, Fisher’s exact test or the chi-squared test was used, as appropriate. The receiver operating characteristic (ROC) curve, sensitivity, specificity, and area under the curve (AUC) were measured to evaluate the levels of laboratory indicators predicting the need for oxygen therapy. We considered that the best cutoff was near the shoulder of the ROC curve because as the sensitivity is progressively increased, there is little or no loss in specificity until extremely high levels of sensitivity are achieved. The cumulative probability that patients with COVID-19 would not require oxygen therapy was described via Kaplan–Meier analysis and analyzed using log-rank test. Patients who required oxygen therapy started this treatment within 7 days after blood sampling, and thus, the analysis was performed within 8 days (follow-up time) after blood sampling.

Multivariate logistic analysis was used to exploit the risk factors for oxygen therapy. P < 0.05 was considered statistically significant. Statistical analysis was performed using BellCurve for Excel by Social Survey Research Information Co. Ltd and EZR by Saitama Medical Center, Jichi Medical University, Saitama, Japan, which is a 120 graphical user interface for R (The R Foundation for Statistical Computing, Vienna, 121 Austria) (Kanda et al., 2013).

## Results

### Characteristics

In total, 287 outpatients who visited Asahikawa City Hospital for the first time were eligible for the study. We excluded five patients who were admitted to other hospitals after they visited our hospital because the clinical course data were missing.

We divided the patients into two groups according to the need for oxygen therapy. S1 Table presents the characteristics of the 282 outpatients included in the study. Among the included patients, 142 received blood tests before any treatment for COVID-19 was started. All blood samples were collected at the outpatient setting. In total, 138 patient samples were collected at the first visit to our hospital’s outpatient unit. Four patient samples were collected at the second or third visit. Table 1 presents the characteristics of 142 patients who underwent blood testing. Details of their comorbidities are presented in S2 Table. Forty patients required oxygen therapy, and the remaining patients recovered without oxygen therapy. The median patient age was 55 (45.5–72.25) years among those requiring oxygen therapy, versus 44.5 (34.3– 52.8) years among those who did not require oxygen therapy (P < 0.001). The proportions of males among patients who did and did not receive oxygen therapy were 57.8% and 57.5% (P > 0.999), respectively. No significant difference was found in BMI, smoking history, receipt of SARS-CoV-2 vaccines, comorbidities, and the time from onset between the patients who did and did not require oxygen therapy. The ROC curves of BMI and age distinguishing the need for oxygen therapy are presented in Fig 1, and the AUCs of age and BMI were 0.699 (0.603–0.795) and 0.604 (0.496–0.713), respectively. The optimal cutoffs of BMI and age were 25.2 kg/m^2^ and 52 years, respectively. The sensitivity and specificity of BMI were 0.611 and 0.619, respectively, and those of age were 0.625 and 0.922, respectively.

**Table 1.** Characteristics of 142 outpatients who underwent blood testing.

|  | No oxygen therapy<br>(n = 102) | Oxygen therapy (n =<br>40) | P |
| --- | --- | --- | --- |
| **Demographics** |  |  |  |
| Male (%) | 59 (57.8%) | 23 (57.5%) | 1 |
| Age (years), median (IQR) | 44.5 (34.3–52.8) | 55 (45.5–72.25) | <0.001* |
| BMI, median (IQR) (missing: 28.7%) | 23.9 (21.5–27.3) | 25.7 (23.1–28.6) | 0.071 |
| Smoking, yes (%) (missing: 33.7%) | 46 (47.4%) | 20 (50%) | 0.852 |
| SARS-CoV-2 vaccines, yes (%) | 4 (3.9%) | 3 (7.5%) | 0.402 |
| Comorbidities (%) | 40 (39.2%) | 21 (52.5%) | 0.211 |
| Time from onset (days), median (IQR) | 5 (3–7) | 6 (3–7.5) | 0.577 |
| **Laboratory findings** |  |  |  |
| WBC (/μL), mean (IQR) | 4550 (3817.5–5460) | 5315 (4157.5–9920) | 0.049* |
| Neutrophil-to-lymphocyte ratio, mean<br>(IQR) | 2.81 (1.94–4.64) | 4.74 (3.87–7.39) | <0.001* |
| AST (U/L), mean (IQR) | 28.5 (22.0–35.75) | 39.0 (28.5–56.5) | <0.001* |
| ALT (U/L), mean (IQR) | 23.0 (15.25–44.5) | 25.5 (17.0–39.25) | 0.462 |
| LDH (U/L), mean (IQR) (missing: 0.7%) | 221 (188–986) | 313 (264–415) | <0.001* |
| Ferritin (ng/mL), mean (IQR) (missing: 14.7%) | 249.7 (105.5–450.4) | 465.8 (214.6–847.0) | 0.004* |
| D-dimer (μg/mL), mean (IQR) (missing: 14.7%) | 0.3 (0.20–0.78) | 1.0 (0.55–1.20) | <0.001* |
| CRP (mg/L), mean (IQR) | 1.96 (0.66–3.21) | 6.29 (3.16–9.75) | <0.001* |

\*Significant at $P < 0.05$

**Figure 1.**
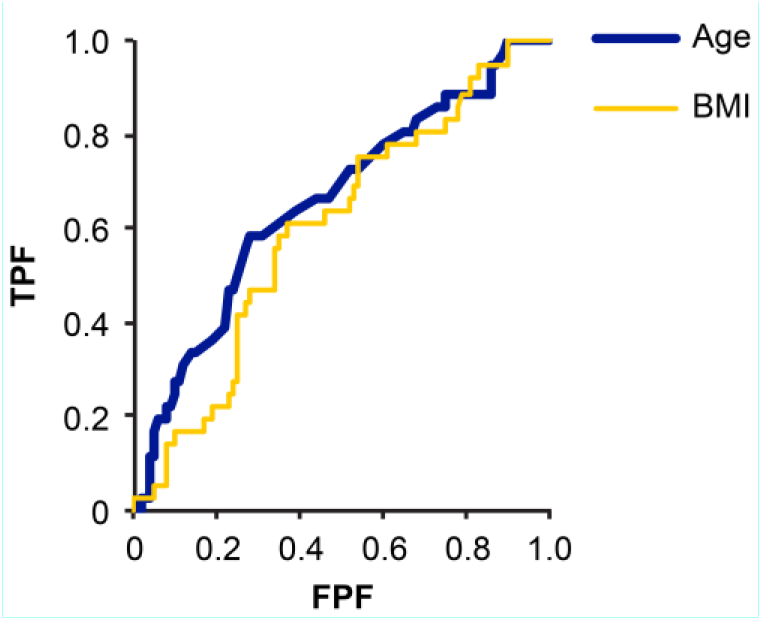
ROC curves of BMI and age distinguishing the need for oxygen therapy.

### Laboratory findings

In patients requiring oxygen therapy, higher WBC (5315/µL [4157.5–9920] vs. 4550/µL [3817.5–5460]/µl, P = 0.049), higher NLR (4.74 [3.87–7.39] vs. 2.81 [1.94–4.64], P < 0.001), higher LDH (313 U/L [264–415] vs. 221 U/L [188–986], P < 0.001), higher CRP (62.9 mg/L [31.6–97.5] vs. 19.6 mg/L [6.6–32.1], P < 0.001), higher ferritin (465.8 ng/mL [214.6–847.0] vs. 249.7 ng/mL [105.5–450.4], P = 0.004), and higher d-dimer (1.0 µg/mL [0.55–1.20] vs. 0.3 µg/mL [0.20–0.78], P < 0.001) were found. ALT levels did not differ between the groups (39.0 U/L [28.55–56.5] vs. 28.5 U/L [22.0–35.75], P = 0.462). We selected WBC, NLR, CRP, AST, and LDH as potential biomarkers predicting the need for oxygen therapy, whereas ferritin and d-dimer were excluded because these data were missing for 14.7% of patients.

The ROC curves of WBC, NLR, CRP, AST, and LDH in distinguishing the need for oxygen therapy are presented in Fig 2. Table 2 presents the AUC, P-value, cutoff, sensitivity, and specificity of each variable. The AUCs of WBC, NLR, CRP, AST, and LDH for predicting the need for oxygen therapy were 0.604, 0.730, 0.799, 0.716, and 0.774, respectively. The optimal cutoffs of WBC, NLR, CRP, AST, and LDH were 5270/µL, 3.546, 36 mg/L, 35 U/mL, and 267 U/mL, respectively. The respective sensitivity and specificity of these variables were as follows: WBC, 0.683 and 0.525; NLR, 0.644 and 0.825; CRP, 0.802 and 0.725; AST, 0.723 and 0.675; and LDH, 0.713 and 0.750. There was no significant difference in AUC among CRP, LDH, NLR, and AST, whereas that of WBC was significantly lower than those of the other variables (P < 0.05). We selected CRP and LDH for further analysis because they had higher AUCs and they are routinely used in daily practice.

**Table 2.** The AUC (95% confidence interval [CI]), P-value, cutoff, sensitivity, and specificity of WBC, NLR, CRP, AST, and LDH in predicting the need for oxygen therapy.

|  | AUC | 95% CI | P-value | Cutoff | Sensitivity | Specificity |
| --- | --- | --- | --- | --- | --- | --- |
| WBC | 0.604 | 0.497–0.711 | 0.056 | 5270/μL | 0.683 | 0.525 |
| NLR | 0.730 | 0.635–0.825 | <0.001* | 3.546 | 0.644 | 0.825 |
| CRP | 0.799 | 0.711–0.886 | <0.001* | 36 mg/L | 0.802 | 0.725 |
| AST | 0.716 | 0.618–0.813 | <0.001* | 35 U/mL | 0.723 | 0.675 |
| LDH | 0.7743 | 0.689–0.856 | <0.001* | 267 U/mL | 0.713 | 0.750 |

**Figure 2.**
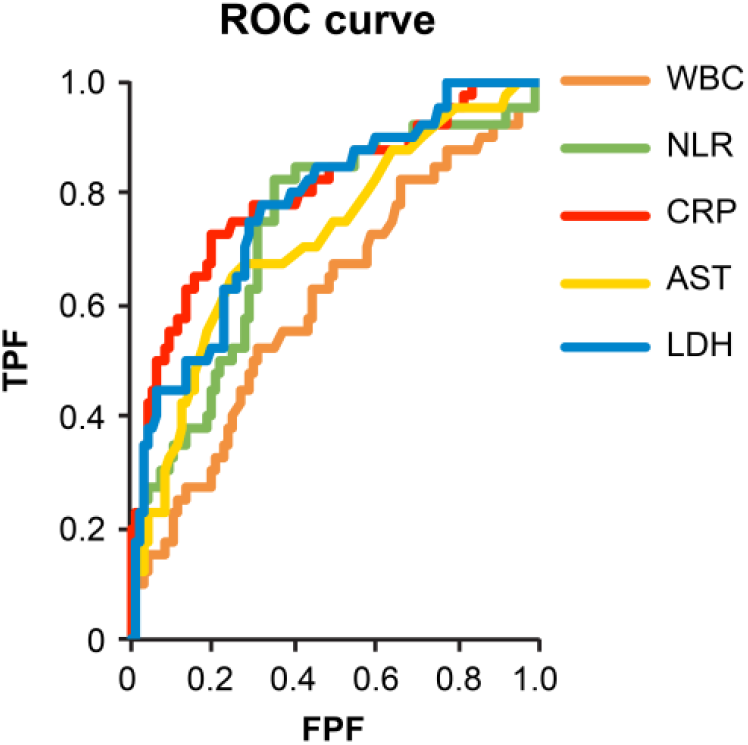
ROC curves of WBC, NLR, CRP, AST, and LDH in distinguishing the need for oxygen therapy.

All patients in the oxygen therapy group started therapy within 7 days of blood testing. The Kaplan–Meier distributions revealed a significant difference in the rates of oxygen therapy between low (<35 mg/L and high (≥35 mg/L) CRP levels (log-rank P < 0.001, Fig. 3) and between low (<267 U/L) and high (≥267 U/L) LDH levels (log-rank P < 0.001*, Fig. 4).

**Figure 3.**
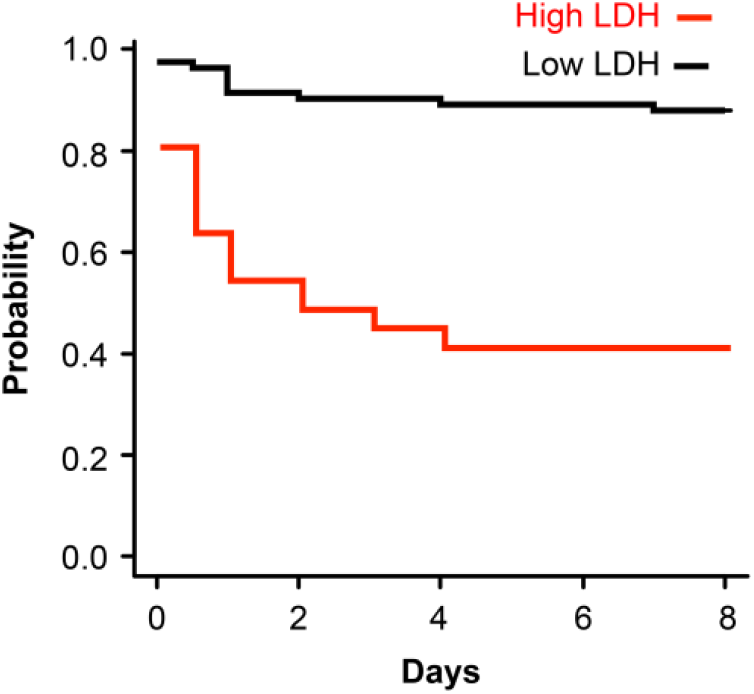
Kaplan–Meier estimates of the cumulative probability that patients with COVID-19 would require oxygen therapy according to plasma CRP < 36 mg/L vs. CRP ≥ 36 mg/L.

**Figure 4.**
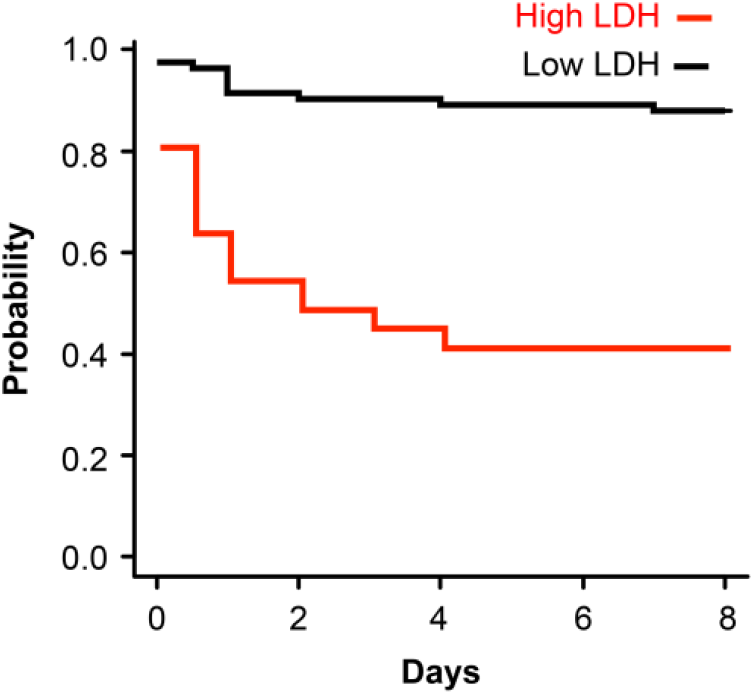
Kaplan–Meier estimates of the cumulative probability that patients with COVID-19 would require oxygen therapy according to plasma LDH < 267 U/L vs. LDH ≥ 267 U/L.

The results of multivariate logistic regression analysis for predicting the need for oxygen therapy according to age, BMI, CRP ≥ 36 mg/L, LDH ≥ 267 U/mL, and comorbidities are presented in Fig 5. Age, BMI, CRP, and LDH were predictive of a significantly higher need for oxygen therapy with odds ratios (95% CIs) of 1.06 (1.02–1.10), 1.15 (1.02–1.29), 7.83 (2.45– 25.1), and 3.9 (1.14–13.3), respectively (Fig 5).

**Fig 5.**
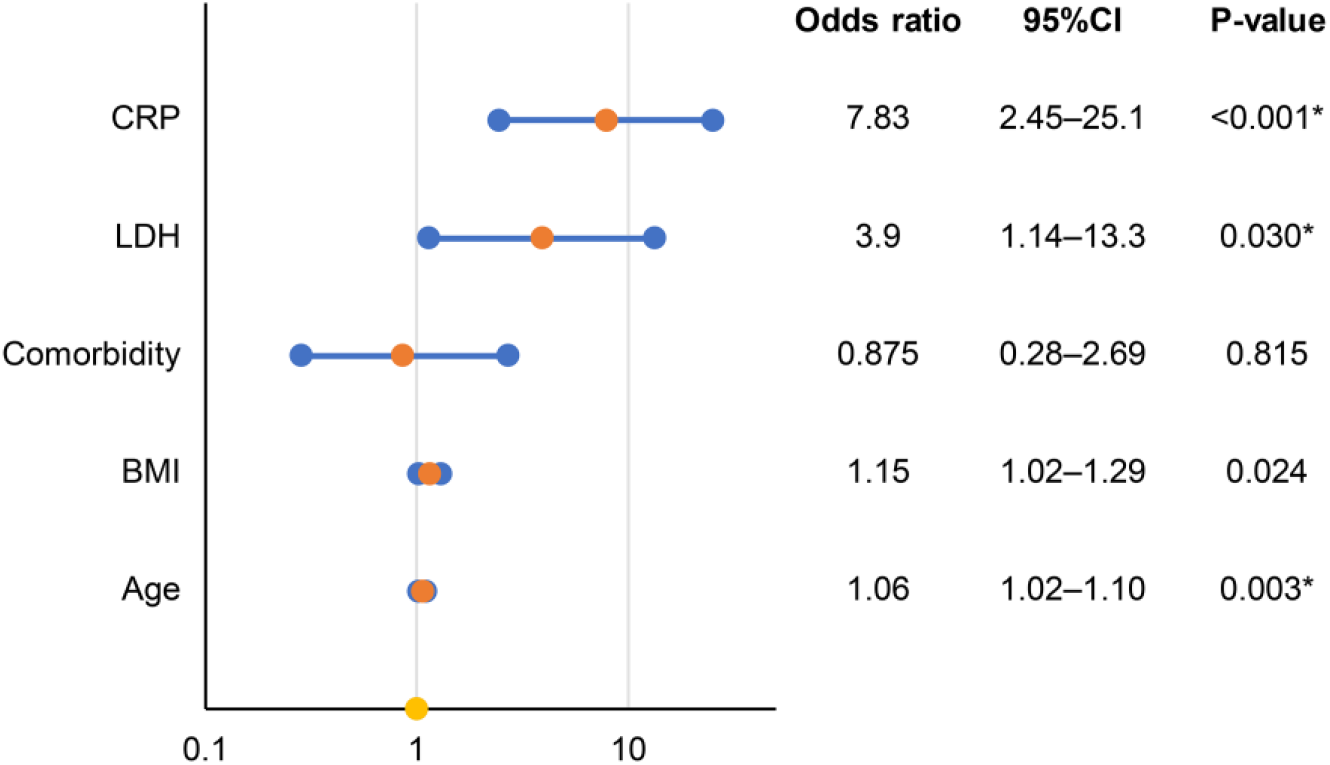
Odds ratios and 95% CIs of age, BMI, CRP, and LDH (multivariate logistic regression analysis)

## Discussion

This study aimed to identify useful laboratory findings predicting the need for oxygen therapy among outpatients with COVID-19. In multivariate analysis, CRP ≥ 36 mg/L, and LDH ≥ 267 U/L are independent risk factors for oxygen therapy. CRP and LDH are widely used and inexpensive to measure. Our findings will help physicians during the management of COVID-19 by indicating the need for frequent follow-up or therapies to avoid poor prognoses.

Excessive inflammation is considered the main cause of critical illness and death in patients with COVID-19 (Luan et al., 2021). CRP levels are obviously increased in the presence of acute inflammation, and they might normalize after inflammation subsides (Luan et al., 2021). CRP levels in the early stage of COVID-19 have been reported to be positively correlated with the presence of lung lesions (Wang et al., 2020). During the process of COVID-19 pneumonia, a cytokine response storm can be triggered, which is associated with high mortality in COVID-19 (Azar et al., 2020). Because the CRP level varies by stage of inflammation, the CRP cutoff predicting severity should change according to the stage. CRP ≥ 110 mg/L is associated with moderate-to-severe acute respiratory distress syndrome in patients with COVID-19 (Poggiali et al., 2020). Patients with CRP > 64.75 mg/L on admission were more likely to have severe complications of COVID-19 (Sadeghi-Haddad-Zavareh et al.,2021). Stringer et al. reported that a CRP cutoff of ≥40 mg/L on admission was a good indicator of mortality in patients with COVID-19 (Stringer et al., 2021). These previous studies focused on CRP levels at admission to predict disease severity. Our study therefore provides useful information by suggesting a CRP cutoff of ≥36 mg/L for identifying outpatients who will require oxygen therapy.

LDH is an inflammatory marker that may serve as a common indicator of acute or severe tissue damage (Sepulveda et al., 2019). Cytokine-mediated lung tissue damage, which is a primary feature of COVID-19, may be responsible for the elevated release of LDH (Martinez-Outschoorn et al., 2011). LDH ≥ 450 U/L on hospitalization was associated with moderate-to-severe acute respiratory distress syndrome in patients with COVID-19 (Poggiali et al., 2020). Li et al. reported that LDH levels of ≥359.5 and ≥277 U/L at admission was associated with death and severe COVID-19, respectively (Li et al., 2020). Although this report used the serum LDH level at admission, the result was similar to those of our study, which suggested that LDH ≥ 267 U/L in outpatients with COVID-19 was predictive of the need for oxygen therapy. Additionally, in multivariate analysis, serum LDH ≥ 267 U/mL remained an independent risk factor for oxygen therapy in our study, in line with the results of Li et al.

Our findings suggest that both CRP ≥ 36 mg/L and LDH ≥ 267 U/L are useful for identifying patients who will require oxygen support in the near future, and there was no statistical difference in the AUC between these variables. We used patients’ samples obtained at their visits, and the samples were obtained a median of 5 days after symptom onset in the nonoxygen therapy group and a median of 6 days in the oxygen therapy group. Meanwhile, patients who needed oxygen support started within 7 days of blood sampling. Because LDH and CRP levels exhibit dynamic changes during the progression of COVID-19, the interval from symptom onset to blood testing should be considered when physicians devise patients’ treatment strategies.

In this study, BMI, age, and comorbidities were also analyzed in a multivariate regression model because they are well-known prognostic factors for COVID-19. The US Food and Drug Administration published criteria for the use of monoclonal antibodies to treat or prevent SARS-CoV-2 in consideration of risk factors including older age (≥65 years), obesity (BMI > 25 kg/m^2^ in adults), pregnancy, and comorbidities such as chronic kidney disease, diabetes, and chronic lung disease (US Food and Drug Administration., 2021). BMI > 25 kg/m^2^ was similar to our cutoff of 25.2 kg/m^2^, but the number the cases was limited in our analysis. However, our analysis assessed all comorbidities collectively because we did not have sufficient numbers of participants to analyze comorbidities separately. The criteria of the Japanese Ministry of Health, Labor and Welfare indicated that patients younger than 50 years are eligible for monoclonal antibody therapy as high-risk patients (Japanese Ministry of Health, Labor and Welfare., 2021). Our age cutoff of 52 years was similar to these criteria.

Regarding study limitations, the number of cases was limited, and this study was performed retrospectively at a single hospital. Only some patients underwent blood testing at their visit based on the judgment of the physicians. Only 4.93% of patients received SARS-CoV-2 vaccines in this study, and the cutoffs might differ according to the receipt of vaccination. Ferritin and d-dimer levels also significantly differed between patients who did and did not receive oxygen therapy; however, the physicians did not measure these parameters in 14.7% of patients. Because all patients were ethnically Japanese, the results should be verified in other populations. Further studies are needed to confirm our findings.

## Conclusions

This study suggests that CRP ≥ 36 mg/L, and LDH ≥ 267 U/L are independent risk factors for oxygen therapy. Further studies concerning biomarkers will help physicians during the management of COVID-19.

## Data Availability

All data produced in the present study are available upon reasonable request to the authors

## Supporting information

**S1 Table. Characteristics of 282 outpatients who visited our hospital during the study period**

**S2 Table. Comorbidities of 142 patients who underwent blood testing**

## Acknowledgements

The authors thank all the participating physicians and other medical staffs for their care of patients with COVID-19.

COVID-19 medical team: Ai Nakamura, Akihito Tampo, Akiko Irie, Akito Uehara, Hidemitsu Sakagami, Kae Takahashi, Kazuki Yamada, Masahide Nakajima, Seisuke Saito, Shin Kukita, Shohei Kuroda, Roku Sato, Yuji Naito, Yuuki Nagasima

