## Supplemental Table 1 for "C-reactive protein and lactate dehydrogenase as prognostic indicators in COVID-2019 outpatients"

**S1 Table. Characteristics of 282 outpatients who visited our hospital during the study period**

| | No oxygen therapy (n = 238) | Oxygen therapy (n = 44) | P |
|---|---|---|---|
| Male (%) | 124 (52.1%) | 25 (56.8%) | 0.680 |
| Age (years), median (IQR) | 38.0 (24.0–48.0) | 55.0 (45.5–73.0) | <0.001* |
| Body mass index, median (IQR) (missing: 28.7%) | 23.8 (20.1–26.2) | 25.7 (22.6–27.6) | 0.045* |
| Smoking, yes (%) (missing: 33.7%) | 75 (36.2%) | 21 (47%) | 0.340 |
| SARS-CoV-2 vaccines, yes (%) | 6 (2.5%) | 3 (6.8%) | 0.151 |
| Comorbidities (%) | 68 (28.6%) | 23 (52.3%) | <0.001* |

IQR, interquartile range; SARS-CoV-2, severe acute respiratory syndrome coronavirus 2
