## Supplemental Table 2 for "C-reactive protein and lactate dehydrogenase as prognostic indicators in COVID-2019 outpatients"

**S2 Table. Comorbidities of 142 patients who underwent blood testing**

| | No oxygen therapy (n = 102) | Oxygen therapy (n = 40) | P |
|---|---|---|---|
| Comorbidities (%) | 40 (39.2%) | 21 (52.5%) | 0.211 |
| COPD (%) | 0 (0%) | 3 (7.5%) | 0.021* |
| Asthma (%) | 7 (6.9%) | 4 (10%) | 0.504 |
| Lung disease excluding COPD and asthma (%) | 5 (4.9%) | 3 (7.5%) | 0.687 |
| Malignancy (%) | 2 (2.0%) | 3 (7.5%) | 0.136 |
| Hypertension (%) | 14 (13.7%) | 6 (15%) | 1 |
| Heart disease excluding hypertension (%) | 5 (4.9%) | 2 (5%) | 1 |
| Renal failure (%) | 1 (1.0%) | 0 (0%) | 1 |
| Diabetes (%) | 14 (13.7%) | 7 (17.5%) | 0.759 |
| Immunosuppressive disease (%) | 2(2.0%) | 0 (0%) | 1 |

COPD, chronic obstructive pulmonary disease
